# Rates and predictors of uptake of formal and informal mental health support during the COVID-19 pandemic: an analysis of 26,740 adults in the UK in lockdown

**DOI:** 10.1101/2021.01.11.21249509

**Authors:** Feifei Bu, Hei Wan Mak, Daisy Fancourt

## Abstract

**Purpose:** The coronavirus disease 2019 (COVID-19) pandemic has put a great strain on people’s mental health. A growing number of studies have shown worsening mental health measures globally during the pandemic. However, there is a lack of empirical study on how people support their mental health during the COVID-19 pandemic. This study aimed to examine a number of formal and informal mental health supports. Further, it explored factors that might be associated with the use of different types mental health support.

**Method:** Data from 26,740 adults in the UCL COVID -19 Social Study were analysed between 13^th^ April, 2020 and 3^rd^ July, 2020. Data were analysed using logistic and Poisson regression models.

**Results:** About 45% of people reported talking to friends or family members to support their mental health, 43% engaging in self-care activities, 20% taking medication, 9% speaking to mental health professionals, 8% talking to a GP or other health professional, and another 8% using helpline or online services. Gender, education, living status, loneliness, pre-existing mental health conditions, general depression and anxiety, coping and personality were found to be associated with the use of mental health support.

**Conclusion:** While the negative impacts caused by the COVID-19 pandemic are inevitable, people can play an active role in managing their mental health. Understanding the patterns and predictors of various kinds of mental health support during the pandemic is crucial for future service planning and delivery through recognising potential barriers to mental health care faced by certain groups.

## Introduction

The stress and anxiety caused by the emergence of the COVID-19 pandemic, the restrictions on social distancing (e.g. quarantine, national lockdown), reduced access to local and health services, changes in working environment and employment schemes (e.g. the furlough scheme), and the closure of leisure events and infrastructures have caused unprecedented impacts on people’s mental health and wellbeing. Multiple studies have highlighted adverse effects on loneliness, stress, anxiety, depression, irritability, confusion, fear, insomnia, guilty and social anxiety (1–4). The proportion of people with a clinically significant level of mental distress increased by around 43%, from 18.9% to 27% during the first UK lockdown (2), with young adults, women, people of lower socio-economic status (SES) and those living with children most negatively affected (2,5,6). Indeed, given the immeasurable cost of personal, social, economic and health burden during COVID-19, it is expected that many countries (including the UK) will face a mental health pandemic that could last for another few years (as has also been shown in other national crisis such as the Great Recession in the US from 2007 to 2009 (7) and other pandemics (4)). As a consequence, protecting mental health has been recognised as high priority to help individuals build their resilience, adapt to inevitable changes caused by the pandemic, cope with adversity, and to prevent the worsening of mental ill health and the experience of suicidal thoughts, self-harm or suicides (8).

In order to promote population mental health, health services across the UK have been providing guidelines and information to help people to get access to social care and support during the pandemic. These include helplines, recommended home workout and relaxation techniques (9). Further, mental health charities, organisations and support groups have been offering formal advice and helpline services to support mental health during the pandemic (e.g. Mental Health Foundation, Samaritans, and YoungMinds). Indeed, a report from Samaritans shows that there was a notable increase in the number of contacts from people who were concerned about COVID-19 and other issues such as finances, social wellbeing and mental health during lockdown in March 2020 (10). Additionally, there have been reports of many individuals drawing on informal support, including using mental health apps, engaging in self-care behaviours, and speaking with friends or family about their mental health. However, a crucial question is whether these services have been reaching those most in need. For instance, it has been shown that individuals from ethnic minority and lower socioeconomic backgrounds experience more barriers (e.g. financial expense, lack of awareness about how to get help, language barriers) to mental health care outside of pandemic situations (11,12). But preliminary research during the pandemic has already suggested that some further high-risk groups are experiencing barriers. For instance, a recent study found that around two in five people engaging in self-harm behaviours and three in five people with self-harm/suicidal thoughts or reporting abuse had not been able to access any type of formal support during lockdown, while nearly half of people who reported abuse, self-harm/suicidal thoughts and self-harm behaviours additionally did not receive any informal support (13). Therefore, it is vital to ascertain in more detail the demographics of people who have not been accessing either formal or informal support in order to inform the targeting of further more specific support towards groups who may be facing more barriers to mental health care.

In light of this, this study used a large sample of adults in the UK to examine how engagement with both formal mental health support (e.g. taking medication, speaking to mental health or other health professionals) and informal mental health support (e.g. helpline or online service, self-care or speaking to family/friends) during COVID-19 varied depending on people’s demographic background, socio-economic characteristics, social factors, mental health, coping strategies, and personality.

## Method

### Participants

Data were drawn from the UCL COVID-19 Social Study; a large panel study of the psychological and social experiences of over 70,000 adults (aged 18+) in the UK during the COVID-19 pandemic. The study commenced on 21st March 2020 involving online weekly data collection from participants for the duration of the COVID-19 pandemic in the UK. Whilst not random, the study has a well-stratified sample that was recruited using three primary approaches. First, snowballing was used, including promoting the study through existing networks and mailing lists (including large databases of adults who had previously consented to be involved in health research across the UK), print and digital media coverage, and social media. Second, more targeted recruitment was undertaken through partnership with recruitment companies focusing on (i) individuals from a low-income background, (ii) individuals with no or few educational qualifications, and (iii) individuals who were unemployed. Third, the study was promoted via partnerships with third sector organisations to vulnerable groups, including adults with pre-existing mental illness, older adults, and carers. The study was approved by the UCL Research Ethics Committee [12467/005] and all participants gave informed consent.

In this study, we used data between 13^th^ April 2020 (when mental health support information started being collected) and 3^rd^ July 2020 (by which point lockdown measures in the UK had been substantially eased but leisure and cultural facilities and community centres remained closed). A total number of 58,260 participants participated at least once during this period. Except for demographic variables collected when participants first joined the study, most other information were collected weekly. However, coping was measured once which was available only for people who participated between 7^th^ and 14^th^ May. After restricting the sample to only participants who provided full information on all variables of interests, we have an analytical sample of 26,740 participants.

### Measures

This study looked at a range of strategies that people used to support their mental health, including 1) taking medication (e.g. anti-depressants), 2) speaking to a mental health professional (e.g. psychiatrist or psychologist), 3) talking to a GP or other health professionals, 4) speaking to someone on a support helpline or accessing online mental health programmes or forums, 5) spending time on self-care activities (e.g. mindfulness, meditation) or using other self-help resource (e.g. books, videos, apps), and 6) speaking to a family member or friend about their mental health. All were coded a binary variable indicating if participants had used any of these strategies at any point between 13^th^ April to 3^rd^ July 2020. Further, we derived a count variable of the total number of different types of mental health support used by each participant.

To understand how different types of mental health support varied across personal characteristics and background, we considered a wide range of potential predictors. These included demographic and socio-economic factors such as age (18-29, 30-45, 46-59, 60+), gender (women vs. men), ethnicity (Black, Asian, and minority ethnic (BAME) vs. white), education (GCSE or below, A levels or equivalent, degree or above), employment status (employed vs. not employed), annual household income (<£30,000 vs. >£30,000) and area of living (rural vs. urban). We also considered social factors, which included living status (living alone, living with others including children, living with others, no child), social network (close friends <3 vs. ≥3) and baseline loneliness level measured by the 3-item UCLA loneliness scale (a short form of the Revised UCLA Loneliness Scale, UCLA-R (14)). Our analysis also adjusted for a set of baseline mental health measures indicating whether participants had any pre-existing diagnosed mental health conditions. These included depression, which was measured by the Patient Health Questionnaire (PHQ-9; a standard instrument for diagnosing depression in primary care (15)), and anxiety measured by the Generalised Anxiety Disorder (GAD-7; a well-validated tool used to screen and diagnose generalised anxiety disorder in clinical practice and research (16)). Both measured at the first week during the observational period. Moreover, we considered psychological factors including personality traits and coping styles. Personality was measured using the short Big Five Inventory (BFI–2) which is comprised of subscales on extraversion, neuroticism, openness, conscientiousness and agreeableness (17). Factor scores for each subscale were derived from confirmatory factor analysis. Coping was measured by the brief COPE Inventory which contains 28 items measuring 14 different types of coping tactics (18). In this study, we derived coping scores based on the four-factor model, including problem-focused (e.g. active coping), emotion-focused (e.g. religion), avoidant (e.g. substance use) and socially supported (e.g. instrumental support) coping strategies (19).

### Analysis

We used logistic regression to calculate the odds ratios (OR) and 95% confidence intervals (CI) that participants used each type of mental health support based on predictor variables. Further, we fitted a Poisson regression model to examine the number of strategies that people used to support their mental health by estimating incidence rate ratios (IRR) and 95% CIs. All data were weighted to the proportions of sex, age, ethnicity, education, and country of living obtained from the Office for National Statistics (20).

In addition to the main analyses, sensitivity analyses were performed using an alternative measure of pre-existing mental health conditions which made use of more specific conditions (e.g. schizophrenia, bipolar disorder, obsessive-compulsive disorder, psychosis etc.) and were available for a reduced number of participants (80%). This yielded largely similar findings (see the Supplement for details). Confirmatory factor analyses were fitted in Mplus V8, but the main analyses were conducted using Stata V15.

## Results

### Descriptive statistics

In our weighted sample, 51% were women and 13% were from Black, Asian or minority ethnic (BAME) backgrounds. On average, 34% of the sample had a degree or other higher qualification, 57% were in employment, 58% with a household income under £30,000, and 21% lived in a rural area. About one in five participants had a pre-existing mental health diagnosis (see Table S1 in the Supplement).

The most commonly used mental health approach during lockdown was talking to friends or family members (45%), followed by engaging in self-care activities (43%) and taking medication (20%). Less common strategies included speaking to mental health professionals (9%), talking to a GP or other health professional (8%), and speaking to someone on a support helpline or using an online programme or forum (8%) (Figure 1a). On average, 37% of participants did not take any action to support their mental health, compared to 24% of people using one strategy, 23% using two strategies, and 17% using three or more strategies to support their mental health (Figure 1b).

**Figure 1.**
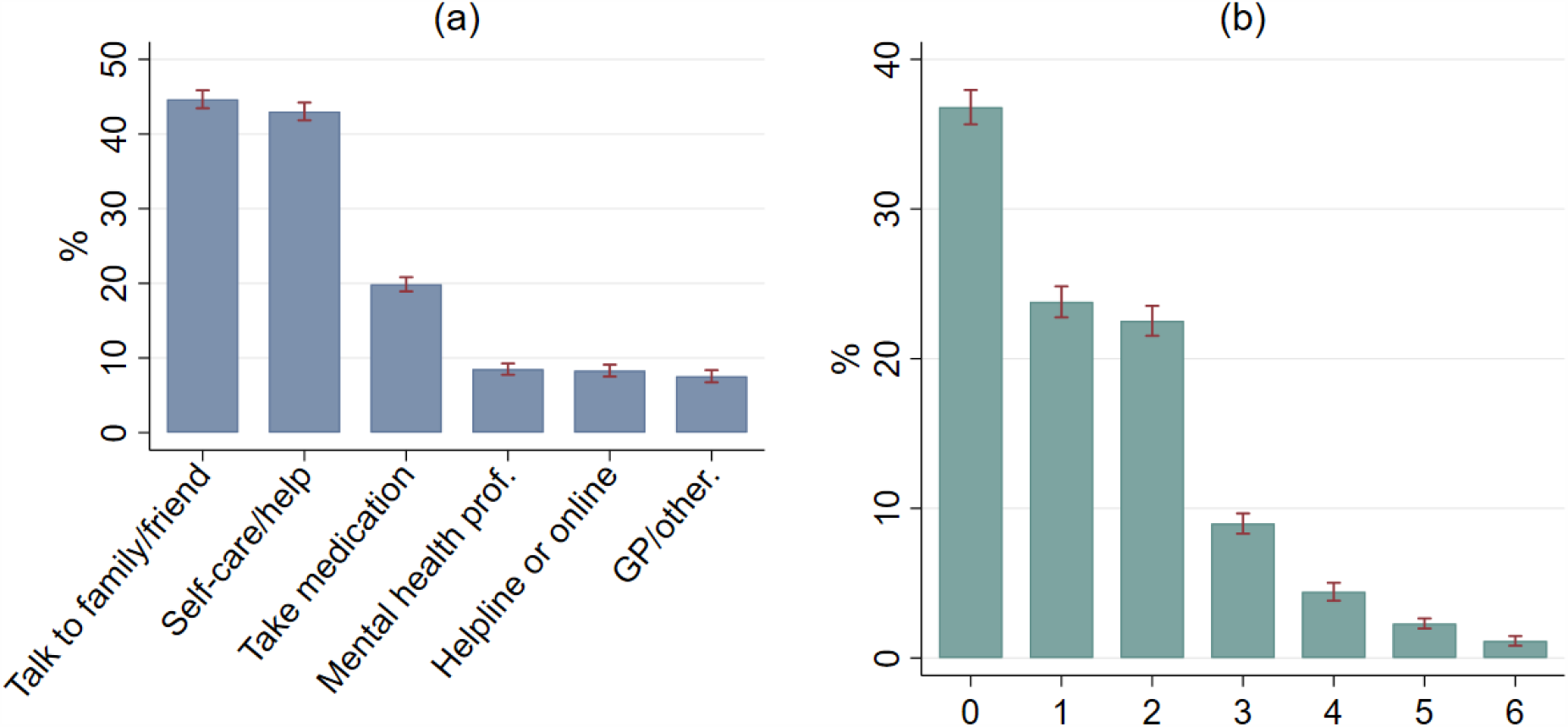
Prevalence of using different types of mental health support

### Demographic and socio-economic characteristics

Comparing to younger adults (aged 18-29), older adults were 2 to 3 times more likely to take medication to support their mental health (OR: 1.92-2.24). However, they were less likely to speak to mental health professionals, seek help through helpline or online forum, engage in self-care activities and speak with friends/family. Women had a 101% higher odds of engaging in self-care activities (OR: 2.01, 95% CI: 1.77-2.27) and a 35% higher odds of speaking to friends/family to support their mental health (OR=1.35, 95% CI: 1.19-1.53). No gender differences were found in terms of using other mental health support. People from BAME groups were less likely to take medication (OR: 0.64 95% CI: 0.42-0.98) but more likely to seek help through helplines or online services (OR: 1.71 95% CI: 1.17-2.48). Ethnicity did not predict use of other types of mental health support. People with higher educational levels were less likely to take medication (OR: 0.76-0.78), but they were more likely to support their mental health through speaking to mental health professionals (OR: 1.42, 95% CI: 1.04-1.94), helpline or online services (OR: 1.38-1.41), self-care activities (OR: 1.30-2.01) and talking to family/friends (OR: 1.25-1.42). People with lower household income had a 25% higher odds of taking medication to support their mental health (OR: 1.25, 95% CI: 1.04-1.50). Those who were employed were more likely to talk to their family/friends to support good mental health (OR: 1.19 95%CI: 1.02-1.38). No association was found between area of living and use of any mental health support (Table 1).

**Table 1.**
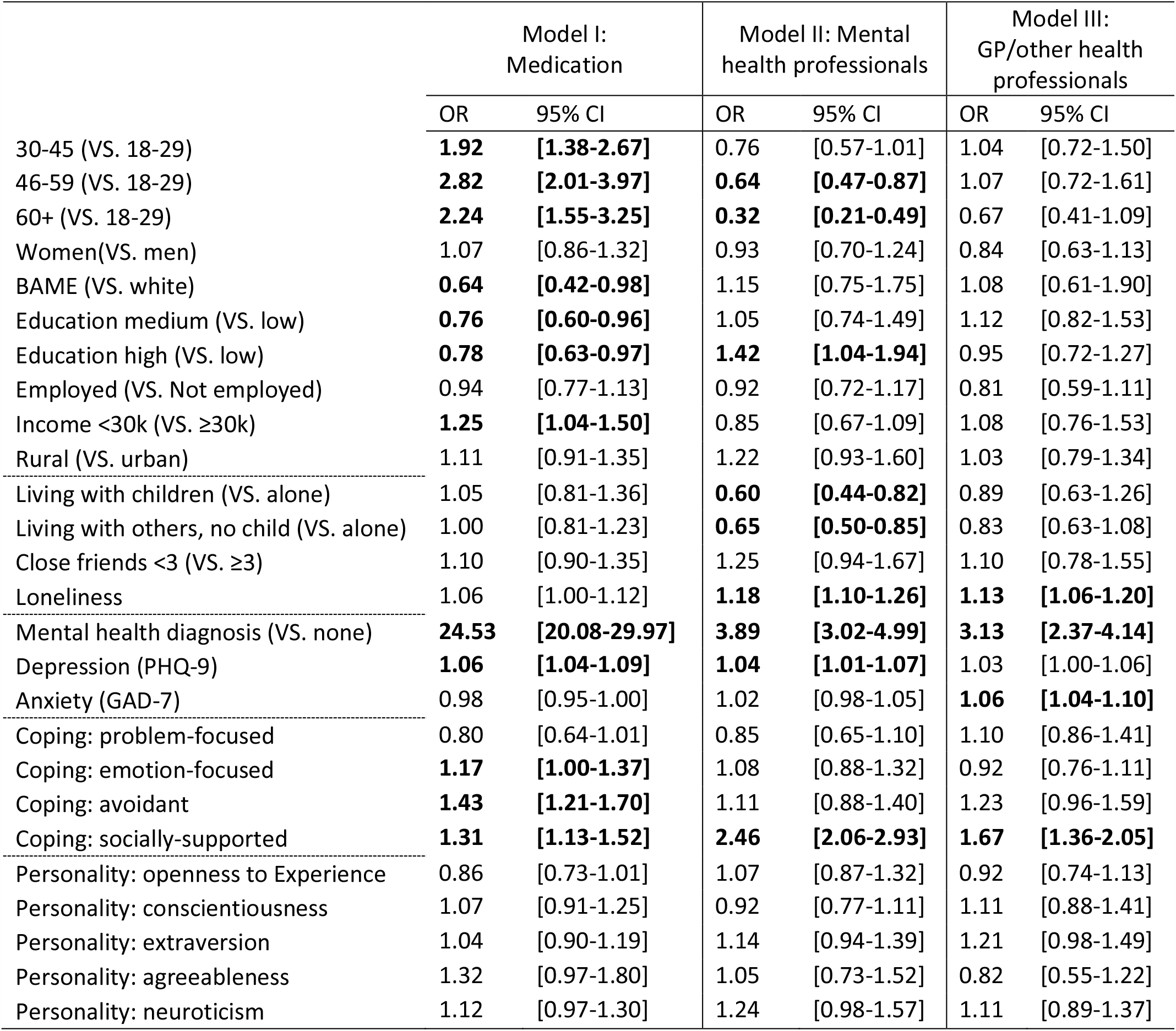
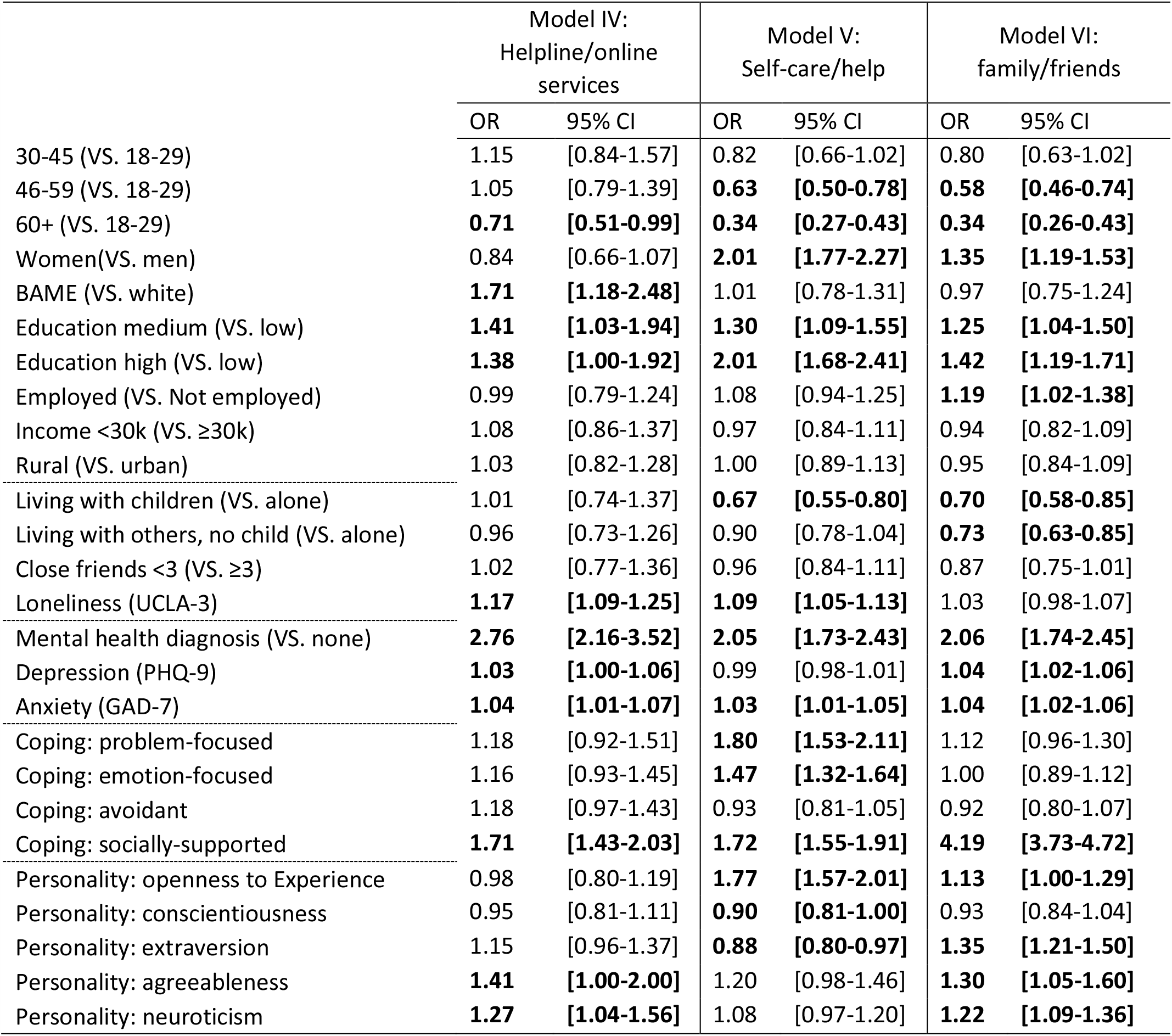
Results from logit model on each strategy to support mental health

|  | Model I:<br>Medication |  | Model II: Mental<br>health professionals |  | Model III:<br>GP/other health<br>professionals |  |
| --- | --- | --- | --- | --- | --- | --- |
|  | OR | 95% CI | OR | 95% CI | OR | 95% CI |
| 30-45 (VS. 18-29) | **1.92** | **[1.38-2.67]** | 0.76 | [0.57-1.01] | 1.04 | [0.72-1.50] |
| 46-59 (VS. 18-29) | **2.82** | **[2.01-3.97]** | **0.64** | **[0.47-0.87]** | 1.07 | [0.72-1.61] |
| 60+ (VS. 18-29) | **2.24** | **[1.55-3.25]** | **0.32** | **[0.21-0.49]** | 0.67 | [0.41-1.09] |
| Women(VS. men) | 1.07 | [0.86-1.32] | 0.93 | [0.70-1.24] | 0.84 | [0.63-1.13] |
| BAME (VS. white) | **0.64** | **[0.42-0.98]** | 1.15 | [0.75-1.75] | 1.08 | [0.61-1.90] |
| Education medium (VS. low) | **0.76** | **[0.60-0.96]** | 1.05 | [0.74-1.49] | 1.12 | [0.82-1.53] |
| Education high (VS. low) | **0.78** | **[0.63-0.97]** | **1.42** | **[1.04-1.94]** | 0.95 | [0.72-1.27] |
| Employed (VS. Not employed) | 0.94 | [0.77-1.13] | 0.92 | [0.72-1.17] | 0.81 | [0.59-1.11] |
| Income <30k (VS. ≥30k) | **1.25** | **[1.04-1.50]** | 0.85 | [0.67-1.09] | 1.08 | [0.76-1.53] |
| Rural (VS. urban) | 1.11 | [0.91-1.35] | 1.22 | [0.93-1.60] | 1.03 | [0.79-1.34] |
| Living with children (VS. alone) | 1.05 | [0.81-1.36] | **0.60** | **[0.44-0.82]** | 0.89 | [0.63-1.26] |
| Living with others, no child (VS. alone) | 1.00 | [0.81-1.23] | **0.65** | **[0.50-0.85]** | 0.83 | [0.63-1.08] |
| Close friends <3 (VS. ≥3) | 1.10 | [0.90-1.35] | 1.25 | [0.94-1.67] | 1.10 | [0.78-1.55] |
| Loneliness | 1.06 | [1.00-1.12] | **1.18** | **[1.10-1.26]** | **1.13** | **[1.06-1.20]** |
| Mental health diagnosis (VS. none) | **24.53** | **[20.08-29.97]** | **3.89** | **[3.02-4.99]** | **3.13** | **[2.37-4.14]** |
| Depression (PHQ-9) | **1.06** | **[1.04-1.09]** | **1.04** | **[1.01-1.07]** | 1.03 | [1.00-1.06] |
| Anxiety (GAD-7) | 0.98 | [0.95-1.00] | 1.02 | [0.98-1.05] | **1.06** | **[1.04-1.10]** |
| Coping: problem-focused | 0.80 | [0.64-1.01] | 0.85 | [0.65-1.10] | 1.10 | [0.86-1.41] |
| Coping: emotion-focused | **1.17** | **[1.00-1.37]** | 1.08 | [0.88-1.32] | 0.92 | [0.76-1.11] |
| Coping: avoidant | **1.43** | **[1.21-1.70]** | 1.11 | [0.88-1.40] | 1.23 | [0.96-1.59] |
| Coping: socially-supported | **1.31** | **[1.13-1.52]** | **2.46** | **[2.06-2.93]** | **1.67** | **[1.36-2.05]** |
| Personality: openness to Experience | 0.86 | [0.73-1.01] | 1.07 | [0.87-1.32] | 0.92 | [0.74-1.13] |
| Personality: conscientiousness | 1.07 | [0.91-1.25] | 0.92 | [0.77-1.11] | 1.11 | [0.88-1.41] |
| Personality: extraversion | 1.04 | [0.90-1.19] | 1.14 | [0.94-1.39] | 1.21 | [0.98-1.49] |
| Personality: agreeableness | 1.32 | [0.97-1.80] | 1.05 | [0.73-1.52] | 0.82 | [0.55-1.22] |
| Personality: neuroticism | 1.12 | [0.97-1.30] | 1.24 | [0.98-1.57] | 1.11 | [0.89-1.37] |

Table 1. Results from logit model on each strategy to support mental health (con't)
|  | Model IV:<br>Helpline/online<br>services |  | Model V:<br>Self-care/help |  | Model VI:<br>family/friends |  |
| --- | --- | --- | --- | --- | --- | --- |
|  | OR | 95% CI | OR | 95% CI | OR | 95% CI |
| 30-45 (VS. 18-29) | 1.15 | [0.84-1.57] | 0.82 | [0.66-1.02] | 0.80 | [0.63-1.02] |
| 46-59 (VS. 18-29) | 1.05 | [0.79-1.39] | **0.63** | **[0.50-0.78]** | **0.58** | **[0.46-0.74]** |
| 60+ (VS. 18-29) | **0.71** | **[0.51-0.99]** | **0.34** | **[0.27-0.43]** | **0.34** | **[0.26-0.43]** |
| Women(VS. men) | 0.84 | [0.66-1.07] | **2.01** | **[1.77-2.27]** | **1.35** | **[1.19-1.53]** |
| BAME (VS. white) | **1.71** | **[1.18-2.48]** | 1.01 | [0.78-1.31] | 0.97 | [0.75-1.24] |
| Education medium (VS. low) | **1.41** | **[1.03-1.94]** | **1.30** | **[1.09-1.55]** | **1.25** | **[1.04-1.50]** |
| Education high (VS. low) | **1.38** | **[1.00-1.92]** | **2.01** | **[1.68-2.41]** | **1.42** | **[1.19-1.71]** |
| Employed (VS. Not employed) | 0.99 | [0.79-1.24] | 1.08 | [0.94-1.25] | **1.19** | **[1.02-1.38]** |
| Income <30k (VS. ≥30k) | 1.08 | [0.86-1.37] | 0.97 | [0.84-1.11] | 0.94 | [0.82-1.09] |
| Rural (VS. urban) | 1.03 | [0.82-1.28] | 1.00 | [0.89-1.13] | 0.95 | [0.84-1.09] |
| Living with children (VS. alone) | 1.01 | [0.74-1.37] | **0.67** | **[0.55-0.80]** | **0.70** | **[0.58-0.85]** |
| Living with others, no child (VS. alone) | 0.96 | [0.73-1.26] | 0.90 | [0.78-1.04] | **0.73** | **[0.63-0.85]** |
| Close friends <3 (VS. ≥3) | 1.02 | [0.77-1.36] | 0.96 | [0.84-1.11] | 0.87 | [0.75-1.01] |
| Loneliness (UCLA-3) | **1.17** | **[1.09-1.25]** | **1.09** | **[1.05-1.13]** | 1.03 | [0.98-1.07] |
| Mental health diagnosis (VS. none) | **2.76** | **[2.16-3.52]** | **2.05** | **[1.73-2.43]** | **2.06** | **[1.74-2.45]** |
| Depression (PHQ-9) | **1.03** | **[1.00-1.06]** | 0.99 | [0.98-1.01] | **1.04** | **[1.02-1.06]** |
| Anxiety (GAD-7) | **1.04** | **[1.01-1.07]** | **1.03** | **[1.01-1.05]** | **1.04** | **[1.02-1.06]** |
| Coping: problem-focused | 1.18 | [0.92-1.51] | **1.80** | **[1.53-2.11]** | 1.12 | [0.96-1.30] |
| Coping: emotion-focused | 1.16 | [0.93-1.45] | **1.47** | **[1.32-1.64]** | 1.00 | [0.89-1.12] |
| Coping: avoidant | 1.18 | [0.97-1.43] | 0.93 | [0.81-1.05] | 0.92 | [0.80-1.07] |
| Coping: socially-supported | **1.71** | **[1.43-2.03]** | **1.72** | **[1.55-1.91]** | **4.19** | **[3.73-4.72]** |
| Personality: openness to Experience | 0.98 | [0.80-1.19] | **1.77** | **[1.57-2.01]** | **1.13** | **[1.00-1.29]** |
| Personality: conscientiousness | 0.95 | [0.81-1.11] | **0.90** | **[0.81-1.00]** | 0.93 | [0.84-1.04] |
| Personality: extraversion | 1.15 | [0.96-1.37] | **0.88** | **[0.80-0.97]** | **1.35** | **[1.21-1.50]** |
| Personality: agreeableness | **1.41** | **[1.00-2.00]** | 1.20 | [0.98-1.46] | **1.30** | **[1.05-1.60]** |
| Personality: neuroticism | **1.27** | **[1.04-1.56]** | 1.08 | [0.97-1.20] | **1.22** | **[1.09-1.36]** |

### Social factors

With respect to social factors, people who lived alone had higher odds of talking to mental health professionals (OR: 1.49-1.67), engaging in self-care activities (OR: 1.49, 95% CI: 1.25-1.82) and talking to family/friends (OR: 1.37-1.43) than those who lived with others. There was no evidence that the size of social network was related to use of mental health support. However, the results show that those who were lonelier were more likely to use most types of mental health support (OR: 1.09-1.18), except for taking medication and talking to family/friends (Table 1).

### Mental health

In relation to mental health, people with pre-existing mental health diagnoses were more likely to use all approaches to support their mental health. In particular, the odds of taking medication for someone with a mental health diagnosis was nearly 25 times of that for those without a diagnosis (OR: 24.53 95% CI: 20.08-29.97). Further, people with a high level of depression were more likely to take medication (OR: 1.06 95% CI: 1.04-1.09) and speak to mental health professionals (OR: 1.04 95% CI: 1.01-1.07), whereas those with a high level of anxiety were more likely to speak to a GP/other health professionals (OR: 1.04 95% CI: 1.02-1.06) and engage in self-care activities (OR: 1.03 95% CI: 1.01-1.05). Higher levels of depression and anxiety were both associated with higher odds of using helpline/online services and talking to family/friends (Table 1).

### Psychological factors

Regarding psychological factors, people with a problem-focused coping strategy were more likely to engage in self-care activity (OR: 1.80 95% CI: 1.53-2.11), whereas those with an avoidant coping strategy were more likely to take medication (OR: 1.43 95%CI: 1.21-1.70). Those with an emotion-focused coping strategy were more likely to support their mental health through medication (OR: 1.17 95%CI: 1.00-1.37) and self-care (OR: 1.47 95%CI: 1.32-1.64). A socially-supported coping strategy was positively associated with all types of mental health support, in particular talking to family/friends (OR: 4.19 95% CI: 3.73-4.72). There was also evidence that personality was associated with using different mental health support. People with high levels of agreeableness and neuroticism were more likely to seek mental health support through helpline/online services (OR: 1.27-1.41) and family/friends (OR: 1.22-1.30). People who were more open to experiences or extraverted were also more likely to seek support from family/friends (OR: 1.13-1.35). However, they differed in that open to experience was associated with higher odds of self-caring (OR: 1.77, 95% CI: 1.57-2.01), but extraversion with lower odds (OR: 0.88, 95% CI: 0.80-0.97) (Table 1).

### Number of types of mental health accessed

When looking at the number of different mental health approaches people used during COVID-19, results show that people aged 60 or above took fewer approaches to support their mental health compared with younger adults (IRR: 0.66 95% CI: 0.62-0.71). In contrast, females (IRR: 1.14 95% CI: 1.09-1.20), people with higher educational levels (IRR: 1.08-1.20), those who lived alone (IRR: 1.09-1.13), those with a higher level of loneliness (IRR: 1.05 95% CI: 1.03-1.06), depression (IRR: 1.01 95% CI: 1.01-1.02) and anxiety (IRR: 1.01 95% CI: 1.00-1.01), and people with a pre-existing mental health diagnosis (IRR: 1.88 95% CI: 1.80-1.97) used more approaches to support their mental health. In addition, people with various coping strategies (IRR: 1.04-1.42) and those with higher levels of openness (IRR: 1.07 95% CI: 1.03-1.11), extraversion (IRR: 1.05 95% CI: 1.10-1.08), agreeableness (IRR: 1.09 95% CI: 1.02-1.17) and neuroticism (IRR: 1.09 95% CI: 1.05-1.13) also used more types of mental health support. Finally, ethnicity, employment status, household income, living area, social network and level of conscientiousness were not related to the number of types of mental health support people accessed (Figure 2).

**Figure 2.**
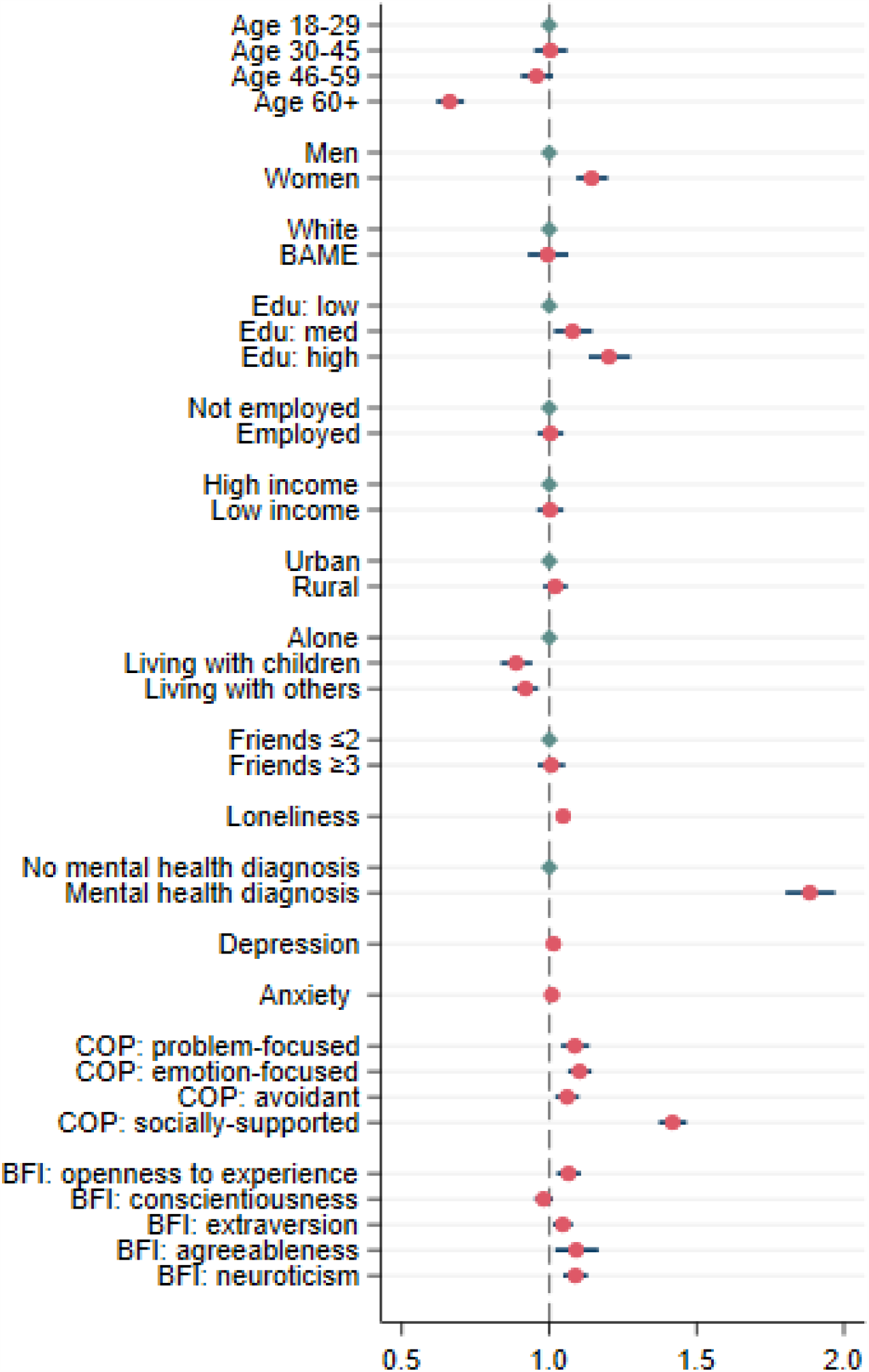
Incident risk ratios and 95% confidence intervals from the Poisson regression model

## Discussion

This study examined what kinds of mental health support individuals accessed during the UK lockdown and how this accessing of support was patterned amongst different demographic groups. Informal mental health support strategies such as talking to family/friends and engaging in self-care activities were the most commonly-used approaches for people to support good mental health during the pandemic, followed by use of medication. On the contrary, formal approaches such as talking to mental health professionals, a GP or other health professionals, and speaking to someone on a support helpline were the least commonly used approaches. Some groups at higher risk of mental ill-health during lockdown did make higher use of support strategies, including women, people living alone, people with a pre-existing mental health diagnosis or people who were experiencing higher levels of anxiety or depression. However, there were also other groups who were at risk of poorer mental health and who also were less likely to access support strategies including individuals with lower educational qualifications.

Some groups such as socially disadvantaged individuals had already been identified as being at a higher risk for experiencing difficulties in accessing mental health services before the pandemic (21). It has been shown that people with lower education level are more likely to encounter acceptability barriers, where they prefer to manage their mental health themselves, do not know how or where to get help, and are afraid to ask for help (21). Compounding these existing inequalities, people from lower socio-economic backgrounds experienced more adversities during the pandemic (e.g. loss of work, unable to pay bills, unable to access sufficient food ad medication) (22) and had poorer trajectories of depression and anxiety (6). As such, it is known that this group was in more need of mental health support. So it is concerning that they had lower levels of engagement with both formal and informal mental health support. This highlights the need for more targeted interventions that aim to reduce the increasing socio-economic inequalities as the pandemic continues. On the contrary, some groups are found to have higher use of support strategies during the pandemic, including women and people living alone. This may explain why these groups, despite having higher levels of depression and anxiety when lockdown commenced, experienced a faster recovery in their mental health during the lockdown period (6).

As well as differences in the overall cumulative number of approaches used, there were also differences in the use of specific mental health approaches amongst different groups. Demographically, while older adults were more likely to take medication to support mental health, younger adults were more likely to use alternative approaches to maintain good mental health (such as talking to mental health professionals, using helpline or online services, self-care activities and talking to family/friends). This difference can be explained by cohort effects, with younger generations more used to communicating their emotions and mental health issues with others in the wake of campaigns such as Mental Health Matters and Heads Together (23,24). In line with previous studies, women were more likely than men to support their mental health via informal strategies, including engaging in self-care activities and sharing their concerns and worries with family/friends (25,26). However, while we found that people with BAME background were less likely to manage their mental health through medication (in line with previous literature which show that the prescription of mental health medication was lower amongst minority ethnic groups (27,28)), they were more likely to use helpline or online services. We also found that people who are socially isolated or lonely, and those who with mental illnesses or symptoms (e.g. depression or anxiety) were more likely to take both formal (e.g. medication) and informal (e.g. speaking to family/friends) mental health approaches. This is in line with previous literature that mental health services are commonly used by people with a mental health diagnosis or those who are struggling with emotional problems (29,30). Finally, individuals with higher educational levels and income were also less likely to use medication to support mental health. This finding corresponds with previous studies conducted before the pandemic, which found that people without qualifications were more likely to use antidepressant drugs and anti-anxiety medication (31,32).

Our study has also shown some further results that have received less attention in previous literature. For example, we found that mental health approaches vary depending on people’s coping strategies when they experience stress, and their personality. Specifically, people who employed problem-focused coping strategies were more likely to engage in self-care activities to manage their mental health during the pandemic, whereas those who employed avoidant coping strategies were more likely to use medication. This could highlight the desire for faster temporary relief from overwhelming emotions amongst people with avoidant coping styles (33), in comparison to a willingness to more proactively seek to control and manage symptoms amongst people with approach-focused coping styles (34). Further, we found that use of formal mental health supports did not vary by personality, but use of informal supports did. Notably, while sharing concerns/worries with family/friends were a common strategy for people of nearly all types of personality (except for conscientiousness), people who were more neurotic tended to seek external help from helpline services or from family/friends. Alternatively, people who were more open were more likely to engage in self-care. The results are supported by a previous meta-analysis study which shows that neuroticism was positively associated with emotional support for coping strategies, whereas high levels of openness were related to problem-solving strategies (35).

This research used a large, well-stratified sample weighted to population proportions to identify the predictors of different approaches people take to support their mental health. However, there are some limitations to the study. Firstly, the data on use of mental health support strategies relied on participants’ self-reports, so could be affected by recall bias or an unwillingness to disclose this information. However, as questions were asked weekly (limiting the length of time people had to remember their actions for) and responses were anonymous, these sources of bias are anticipated to be small. Secondly, our study asked participants about six different types of formal and informal mental health support strategies, but it is possible that participants might have also been using alternative approaches to manage their mental health not captured in our questions, including risky behaviours such as substance use. In addition, data on participants’ previous use of mental health services prior to the pandemic are not available, so it is not known whether participants started using specific strategies during the pandemic or whether these were simply a continuation of previous habits. Further, in line with previous studies (36), we found that people from ethnic minority backgrounds were less likely to access structured mental health services. However, due to limitations in statistical power, we were only able to explore ethnicity as a binary in these analyses, but recognise that such simple categorisation likely misses nuances of experience in specific ethnic groups. Further, it remains unclear what the barriers to accessing services were amongst people from BAME backgrounds (e.g. language barriers, discrimination or social stigma) (37–39). Finally, our analyses did not include details of specific psychiatric diagnosis. Further work is needed to understand how service usage varied for specific clinical groups.

Given limited resources in mental health services and given the growing mental health problems during and after the pandemic, it is important to understand what approaches people employ to support good mental health and to identify which groups are not receiving adequate mental health support. The results of this study suggest that certain groups require more specific mental health support, in particular people with higher levels of loneliness, depression and anxiety during the pandemic and people with a diagnosed mental health condition for which they may have been receiving support before the pandemic. It is promising that talking to family/friends about mental health and self-care activities were the most commonly used strategies during lockdown as it suggests a recognition of the challenges to mental health posed by the pandemic and an openness in discussing mental health issues. However, such strategies may be insufficient for the management of more severe types of mental illness and this, combined with the identification of groups facing more barriers to mental health support, suggests the importance of developing more specific policies and programmes as the pandemic continues.

## Data Availability

Anonymous data will be made available following the end of the pandemic.

## Declarations

### Ethics approval and consent to participate

Ethical approval for the COVID-19 Social Study was granted by the UCL Ethics Committee. All participants provided fully informed consent. The study is GDPR compliant.

### Availability of data and material

Anonymous data will be made available following the end of the pandemic.

### Code availability

Stata syntax are available from the authors upon request.

### Authors’ contributions

DF and FB conceived and designed the study. FB analysed the data and FB, HWM and DF wrote the first draft. All authors provided critical revisions. All authors read and approved the submitted manuscript.

## Supplementary material

**Table S1.** Sample characteristics

|  | Percentage/<br>Mean (standard deviation) |
| --- | --- |
| Age: 18-29 | 19.47% |
| Age: 30-45 | 26.06% |
| Age: 46-59 | 24.14% |
| Age: 60+ | 30.33% |
| Women(VS. men) | 50.68% |
| BAME (VS. white) | 12.78% |
| Education: low | 32.58% |
| Education: medium | 33.81% |
| Education: high | 33.61% |
| Employed (VS. Not employed) | 57.34% |
| Income <30k (VS. ≥30k) | 47.68% |
| Rural (VS. urban) | 20.58% |
| Living alone | 18.98% |
| Living with children | 24.36% |
| Living with others, no child | 56.66% |
| Close friends <3 (VS. ≥3) | 30.60% |
| Loneliness | 4.98 (1.99) |
| Mental health diagnosis (VS. none) | 20.19% |
| Depression (PHQ-9) | 6.85 (6.18) |
| Anxiety (GAD-7) | 5.15 (5.44) |
| Coping: problem-focused | 0.01 (0.50) |
| Coping: emotion-focused | 0.01 (0.66) |
| Coping: avoidant | 0.04 (0.55) |
| Coping: socially-supported | 0.02 (0.69) |
| Personality: openness to Experience | 0.00 (0.58) |
| Personality: conscientiousness | -0.01 (0.71) |
| Personality: extraversion | 0.00 (0.71) |
| Personality: agreeableness | -0.01 (0.37) |
| Personality: neuroticism | 0.00 (0.70) |

**Table S2.**
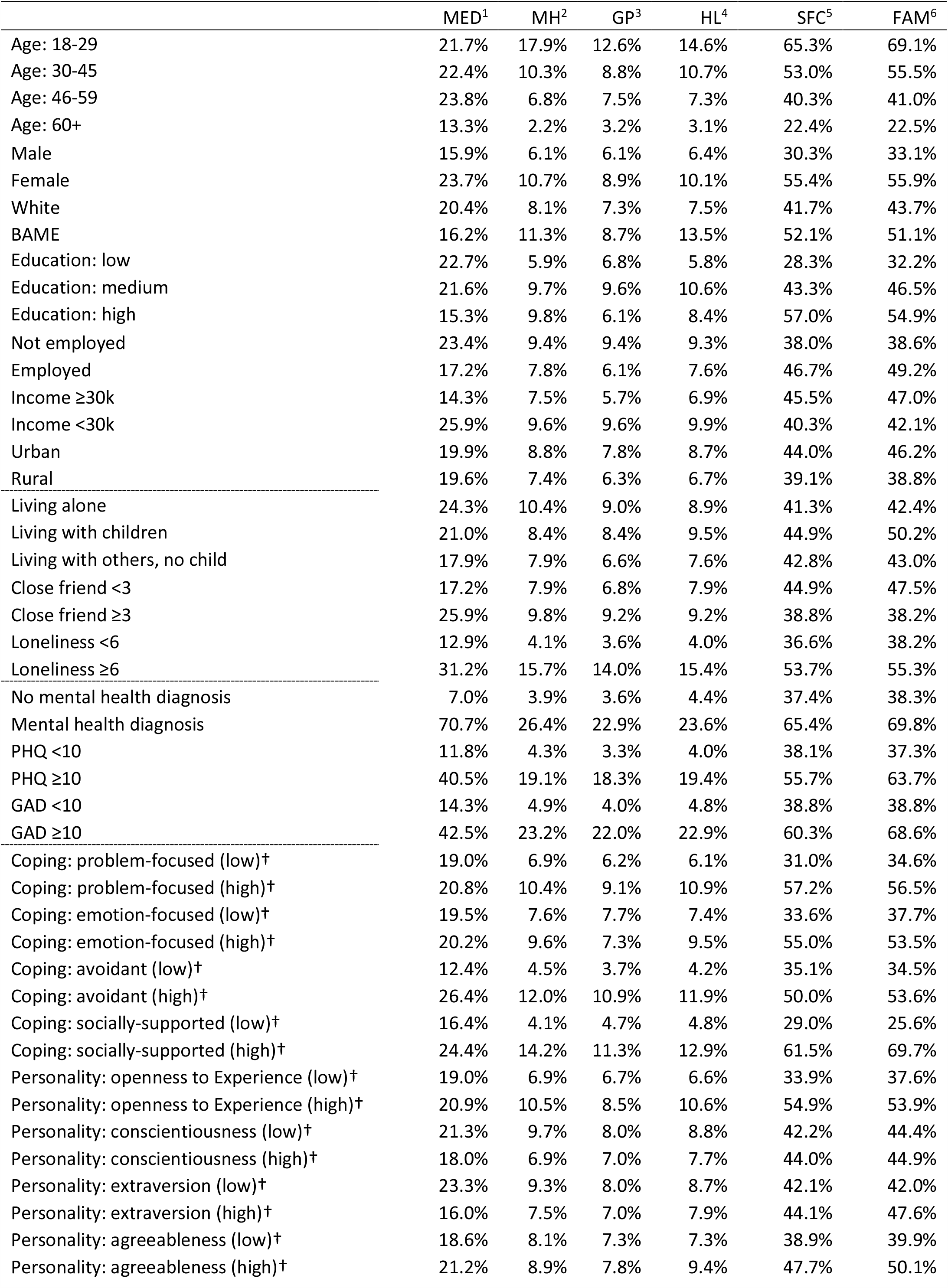
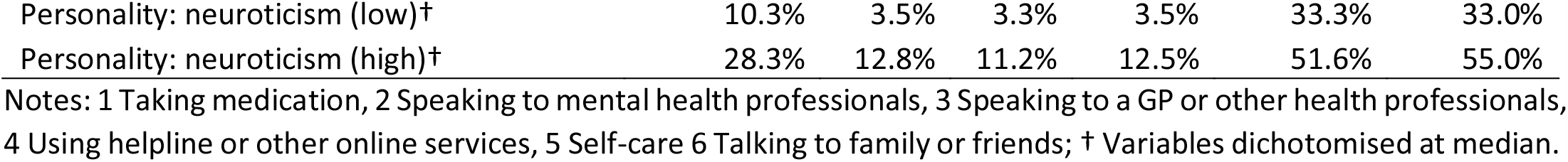
Percentages of people using each strategy by characteristic groups (weighted)

**Table S3.** Frequencies of specific mental health conditions included in the alternative mental health diagnoses measure

| Diagnosis | N |
| --- | --- |
| Schizophrenia | 21 |
| Bipolar disorder | 148 |
| Manic depression | 48 |
| Obsessive-compulsive disorder | 215 |
| Post-traumatic stress disorder | 512 |
| Eating disorder | 222 |
| Psychosis | 31 |
| Panic disorder | 290 |
| Anxiety | 3,370 |
| Depression | 3,112 |
| Other mental health condition | 514 |
| Total number of conditions | 8,483 |
| Total number of participants | 4,864 |

**Table S4.** Cross-tabulation of two mental health measures

| Mental health diagnosis<br>(original) | Mental health diagnosis<br>(alternative) |  |  |
| --- | --- | --- | --- |
|  | No | Yes | Total |
| No | 16,160<br>90.5% | 1,699<br>9.5% | 17,859<br>83.6% |
| Yes | 344<br>9.8% | 3,165<br>90.2% | 3,509<br>16.4% |
| Total | 16,504<br>77.2% | 4,864<br>22.8% | 21,368<br>100.0% |

**Table S5.** Results from logit model on each strategy to support mental health (using alternative mental health diagnosis measure)

|  | Model I:<br>Medication |  | Model II: Mental<br>health professionals |  | Model III:<br>GP/other health<br>professionals |  |
| --- | --- | --- | --- | --- | --- | --- |
|  | OR | 95% CI | OR | 95% CI | OR | 95% CI |
| 30-45 (VS. 18-29) | **2.26** | **[1.55,3.30]** | 0.76 | [0.53,1.10] | 1.08 | [0.72,1.63] |
| 46-59 (VS. 18-29) | **2.84** | **[1.95,4.14]** | **0.64** | **[0.44,0.91]** | 0.97 | [0.65,1.45] |
| 60+ (VS. 18-29) | **2.52** | **[1.68,3.77]** | **0.31** | **[0.20,0.48]** | 0.75 | [0.48,1.16] |
| Women(VS. men) | 1.09 | [0.87,1.37] | 0.95 | [0.68,1.31] | 0.88 | [0.65,1.18] |
| BAME (VS. white) | 0.63 | [0.39,1.00] | 1.18 | [0.69,2.01] | 1.01 | [0.65,1.59] |
| Education medium (VS. low) | 0.80 | [0.64,1.02] | 1.16 | [0.80,1.69] | 1.02 | [0.74,1.40] |
| Education high (VS. low) | **0.78** | **[0.62,0.99]** | **1.67** | **[1.19,2.34]** | 0.94 | [0.68,1.29] |
| Employed (VS. Not employed) | **0.78** | **[0.63,0.96]** | 0.89 | [0.66,1.19] | 0.82 | [0.61,1.11] |
| Income <30k (VS. ≥30k) | 1.05 | [0.85,1.30] | 0.77 | [0.58,1.02] | 1.08 | [0.80,1.46] |
| Rural (VS. urban) | 1.09 | [0.90,1.32] | 1.13 | [0.84,1.53] | 1.07 | [0.80,1.42] |
| Living with children (VS. alone) | 1.00 | [0.74,1.35] | **0.66** | **[0.46,0.95]** | 1.14 | [0.78,1.65] |
| Living with others, no child (VS. alone) | 0.85 | [0.68,1.07] | **0.58** | **[0.43,0.78]** | 0.88 | [0.66,1.18] |
| Close friends <3 (VS. ≥3) | **1.26** | **[1.03,1.56]** | 1.18 | [0.86,1.60] | 0.79 | [0.60,1.04] |
| Loneliness | 1.00 | [0.94,1.07] | **1.17** | **[1.08,1.27]** | **1.19** | **[1.10,1.28]** |
| Mental health diagnosis (VS. none) | **23.68** | **[18.82,29.81]** | **5.99** | **[4.50,7.97]** | **4.67** | **[3.42,6.38]** |
| Depression (PHQ-9) | **1.08** | **[1.05,1.11]** | **1.03** | **[1.00,1.07]** | 1.03 | [1.00,1.07] |
| Anxiety (GAD-7) | 0.97 | [0.94,1.00] | 1.03 | [0.99,1.07] | **1.05** | **[1.02,1.09]** |
| Coping: problem-focused | **0.78** | **[0.62,0.97]** | 0.88 | [0.67,1.16] | 1.13 | [0.87,1.47] |
| Coping: emotion-focused | 1.08 | [0.92,1.27] | 1.16 | [0.93,1.46] | 1.02 | [0.82,1.26] |
| Coping: avoidant | **1.30** | **[1.07,1.57]** | 1.06 | [0.83,1.36] | **1.30** | **[1.04,1.63]** |
| Coping: socially-supported | **1.34** | **[1.15,1.57]** | **2.14** | **[1.75,2.60]** | **1.66** | **[1.37,2.02]** |
| Personality: openness to Experience | 0.95 | [0.78,1.17] | 1.02 | [0.81,1.29] | 1.04 | [0.81,1.33] |
| Personality: conscientiousness | 1.05 | [0.89,1.24] | 0.94 | [0.77,1.15] | 1.05 | [0.86,1.28] |
| Personality: extraversion | 0.90 | [0.77,1.05] | 1.05 | [0.84,1.31] | 0.96 | [0.78,1.19] |
| Personality: agreeableness | 1.31 | [0.96,1.80] | 1.01 | [0.67,1.51] | 1.04 | [0.70,1.55] |
| Personality: neuroticism | 1.07 | [0.90,1.26] | 1.06 | [0.84,1.34] | 1.10 | [0.89,1.36] |

Table S5. Results from logit model on each strategy to support mental health (using alternative mental health diagnosis measure) (con't)
|  | Model IV:<br>Helpline/online<br>services |  | Model V:<br>Self-care/help |  | Model VI:<br>family/friends |  |
| --- | --- | --- | --- | --- | --- | --- |
|  | OR | 95% CI | OR | 95% CI | OR | 95% CI |
| 30-45 (VS. 18-29) | 1.21 | [0.83,1.75] | 0.83 | [0.63,1.09] | 0.80 | [0.58,1.09] |
| 46-59 (VS. 18-29) | 1.04 | [0.74,1.46] | **0.60** | **[0.46,0.78]** | **0.54** | **[0.40,0.74]** |
| 60+ (VS. 18-29) | 0.71 | [0.49,1.03] | **0.31** | **[0.23,0.41]** | **0.31** | **[0.22,0.42]** |
| Women(VS. men) | 0.82 | [0.62,1.08] | **1.96** | **[1.71,2.26]** | **1.41** | **[1.22,1.62]** |
| BAME (VS. white) | 1.43 | [0.94,2.18] | 1.14 | [0.82,1.59] | 1.11 | [0.83,1.50] |
| Education medium (VS. low) | **1.45** | **[1.05,1.99]** | **1.37** | **[1.13,1.66]** | **1.32** | **[1.09,1.61]** |
| Education high (VS. low) | **1.56** | **[1.15,2.11]** | **2.03** | **[1.67,2.47]** | **1.50** | **[1.24,1.82]** |
| Employed (VS. Not employed) | 1.06 | [0.81,1.37] | 1.01 | [0.86,1.18] | **1.26** | **[1.06,1.49]** |
| Income <30k (VS. ≥30k) | 1.18 | [0.91,1.52] | 1.00 | [0.85,1.17] | 0.99 | [0.84,1.17] |
| Rural (VS. urban) | 0.99 | [0.77,1.27] | 1.03 | [0.90,1.18] | 1.05 | [0.91,1.20] |
| Living with children (VS. alone) | 1.08 | [0.74,1.56] | **0.66** | **[0.54,0.82]** | **0.64** | **[0.52,0.79]** |
| Living with others, no child (VS. alone) | 0.91 | [0.69,1.20] | 0.89 | [0.76,1.05] | **0.72** | **[0.61,0.85]** |
| Close friends <3 (VS. ≥3) | 0.97 | [0.71,1.32] | 0.98 | [0.84,1.14] | 0.89 | [0.76,1.06] |
| Loneliness (UCLA-3) | **1.15** | **[1.08,1.24]** | **1.09** | **[1.04,1.14]** | 1.01 | [0.96,1.06] |
| Mental health diagnosis (VS. none) | **3.53** | **[2.68,4.64]** | **1.78** | **[1.50,2.11]** | **2.02** | **[1.69,2.42]** |
| Depression (PHQ-9) | **1.04** | **[1.01,1.07]** | 1.01 | [0.99,1.03] | **1.04** | **[1.01,1.06]** |
| Anxiety (GAD-7) | **1.06** | **[1.02,1.09]** | **1.03** | **[1.00,1.05]** | **1.05** | **[1.02,1.07]** |
| Coping: problem-focused | **1.32** | **[1.02,1.72]** | **1.98** | **[1.65,2.39]** | 1.10 | [0.92,1.30] |
| Coping: emotion-focused | 1.21 | [0.99,1.48] | **1.49** | **[1.31,1.69]** | 0.99 | [0.87,1.13] |
| Coping: avoidant | 1.07 | [0.86,1.33] | 0.91 | [0.78,1.05] | 0.95 | [0.80,1.12] |
| Coping: socially-supported | **1.58** | **[1.30,1.92]** | **1.71** | **[1.52,1.92]** | **4.18** | **[3.66,4.78]** |
| Personality: openness to Experience | 0.99 | [0.79,1.24] | **1.65** | **[1.43,1.90]** | 1.06 | [0.91,1.23] |
| Personality: conscientiousness | 0.86 | [0.72,1.04] | 0.92 | [0.81,1.04] | 0.95 | [0.84,1.08] |
| Personality: extraversion | 1.08 | [0.88,1.32] | 0.92 | [0.83,1.03] | **1.31** | **[1.16,1.48]** |
| Personality: agreeableness | **1.54** | **[1.03,2.29]** | 1.17 | [0.93,1.48] | **1.34** | **[1.04,1.71]** |
| Personality: neuroticism | 0.96 | [0.77,1.20] | 1.08 | [0.95,1.22] | **1.17** | **[1.03,1.33]** |

